# Nurse staffing levels and patient outcomes: a systematic review of longitudinal studies

**DOI:** 10.1101/2021.09.17.21263699

**Authors:** Chiara Dall’Ora, Christina Saville, Bruna Rubbo, Lesley Y Turner, Jeremy Jones, Peter Griffiths

## Abstract

**Background:** The contribution of registered nurses (RN) towards safe patient care has been demonstrated by several studies. However, most of the evidence is cross-sectional, hence the inability to demonstrate that staffing levels precede patient outcomes. No reviews have summarised longitudinal studies considering nurse staffing and patient outcomes.

**Objectives:** To synthesise longitudinal studies focusing on associations between nurse staffing levels and patient outcomes.

**Methods:** Systematic review. We conducted our search in 2020 and updated it in July 2021. We searched Medline, CINAHL, Embase and the Cochrane Library. We used the ROBINS-I tool for assessing risk of bias. We reported results narratively grouped by outcome.

**Results:** 28 papers were included. Most studies were either at serious (n=12) or critical (n=6) risk of bias, with 3 studies at low risk of bias. Studies were conducted in a variety of settings and populations. Notwithstanding the limitations, findings are consistent with an overall picture of a beneficial effect from higher RN staffing on preventing patient death. Studies with the greatest risk of bias were judged as most likely to underestimate the effect of higher RN staffing. The evidence is less clear for other patient outcomes, but estimates, though at moderate or serious risk of bias, indicate that higher RN staffing is likely to lead to better patient outcomes. Evidence about the contribution of other nursing staff groups and skill mix of the team is unclear.

**Conclusion:** There is a likely causal relationship between low RN staffing and harm to patients, although uncertainties remain regarding the magnitude of effect. To address these uncertainties, future studies should be conducted in more than one hospital and using standardised measures when reporting staffing levels.

## INTRODUCTION

Nursing staff account for nearly half of the health workforce worldwide, and the cost of providing inpatient hospital nursing is one of the main cost drivers for health systems.^1^ Providing nurse staffing levels that match patient demand is key to deliver cost-effective health services. In the face of increasing financial pressures and budget constraints, registered nurses (RNs) are sometimes viewed simply as a costly labour input, which can often be substituted with lower paid unregistered staff.^2^ This has raised questions about the contribution of RNs and other nurses in the workforce in ensuring patients receive safe and high-quality care, including preventing deterioration and death among hospital inpatients. Besides RNs, the nursing workforce comprises of unregistered nursing assistants (NAs), and licenced practical nurses/nursing associates (LPNs), who accessed the profession not through a university degree, but rather by completing a formal training programme.

When considering the associations between nurse staffing and patient outcomes in inpatient hospital settings, the breadth of the evidence is apparent, with hundreds of studies published to date, most of which support a conclusion that the higher the RN staffing, the lower the rate of adverse patient outcomes, including death.^3^ The volume of the evidence has led many to question whether more studies are needed because the implications for policy and practice are clear and the evidence definitive.^4^ ^5^ However past reviews of this literature,^6-9^ have noted the preponderance of cross-sectional studies. Such studies are unable to establish that the observed variation in staffing levels, typically measured at a hospital level average, was experienced by the patients whose outcomes were measured, typically aggregated at the hospital level over a year. This approach cannot accurately estimate the nursing care available to individual patients whose outcomes are studied.

The limitations of the evidence are such that for some, a causal relationship is still questionable, although a careful analysis applying epidemiological principles suggests that the body of evidence is consistent with a “cause and effect” relationship.^3^ ^6^ Nonetheless, the indirect associations reported in most studies means it is impossible to estimate the effect of change in staffing levels without bias.

In recent years, there has been a steady increase in studies using routinely collected data in healthcare, including studies that use electronic rostering systems and patient records to link patients to the staffing levels they are exposed to throughout their hospital stay.^10^ ^11^ Such studies have the potential to establish the staffing levels that individual patients have been exposed to *prior* to experiencing the outcome and so directly explore the effect of variation. In addition to removing many potential sources of bias associated with cross-sectional studies, longitudinal studies avoid the intrinsic limitation of cross-sectional studies, as the presumed cause can be shown to precede the outcome of interest. There is no summary of studies using longitudinal designs to explore the impact of staffing levels on patient and organisational outcomes in inpatient hospital settings. Therefore, the aim of this systematic review is to synthesise the evidence on the effect of exposure to variation in nurse staffing levels on subsequent patient outcomes.

## METHODS

### Eligibility criteria

We included all quantitative, longitudinal studies that measured nurse staffing and assessed the association between variation in staffing and subsequent patient outcome(s). We included prospective, retrospective, cohort, case-control, randomised or quasi-randomised controlled trials, controlled before-and-after studies, interrupted time series, difference-in-difference or panel studies. To be eligible, studies must establish a temporal link between nurse staffing and patient outcomes, such that variation in nurse staffing levels preceded the outcomes patients experienced. Studies with repeated measures which deployed a cross-sectional analysis were excluded except where difference in difference designs were used to show an association between change in staffing and change in outcomes over time. Similarly, we included studies where planned interventions were implemented and natural experiments where the effect of an exogenous ‘shock’ (e.g. changes in legislation or major policies designed to alter staffing levels) involving a change in staffing was studied. We did not specify a list of patient outcomes *a priori*, but we defined patient outcomes as any outcomes experienced by patients as opposed to staff, families, and the healthcare systems.

### Study selection and data extraction

We registered our review protocol in Prospero (Blinded_for_peer_review). We conducted our search in October 2020 and updated it in July 2021. We searched Medline, CINAHL, Embase and the Cochrane Library. The complete search strategy can be found in Appendix 1.

One reviewer (XXX) undertook the first screening to remove duplicates and irrelevant studies. Potentially relevant papers were then further screened with a more detailed assessment of titles and abstracts. At this stage, all other reviewers assessed samples of 10 studies each to ensure that there was consistency, and to identify points of ambiguity and uncertainty in selection criteria. Full texts of studies that remained after this screening were retrieved and detailed assessment was made against the criteria. All full text papers were assessed by XXX and another reviewer, and any disagreements were resolved by discussion and reaching consensus among all reviewers.

From each included study we extracted author(s), year; country and setting; sample size; measure(s) of staffing levels; outcomes and risk-adjustment; findings.

### Assessment of risk of bias

We used the ROBINS-I tool for assessing risk of bias in non-randomised studies; this tool focuses on a study’s internal validity, and bias is defined as a tendency for study results to differ systematically from the results expected from a randomised trial, conducted on the same participants and with no flaws in its conduct.^12^ When using the tool to assess studies of natural variation in staffing we interpreted “intervention” as variation in exposure to nurse staffing levels. The risk of bias due to different methodological aspects (called “domains”) was assessed: confounding, selection of participants into the study, classification of interventions, deviation from intended interventions, missing data, measurement of outcomes, and selection of the reported results. Each domain of the ROBINS-I tool was graded as either low, moderate, serious, or critical risk of bias, and the domain grade with the highest risk of bias determined the overall risk of bias grade. According to the ROBINS-I guidance,^12^ we excluded studies at critical risk of bias from our synthesis,^13-18^ but these studies’ characteristics are reported in Appendix 2.

We considered whether bias was likely to over or under estimate nurse staffing effects. Two reviewers independently assessed the risk of bias for each study, and where there was disagreement, this was resolved by collective discussion.

### Evidence synthesis

We were unable to identify groups of studies that used measures of staffing and outcomes that were sufficiently comparable to pool in a statistical meta-analysis. Therefore, we performed a narrative synthesis with results grouped by outcome. Where studies performed more than one analysis, we reported results at the analysis level rather than summarising them at the study level. Outcome measures were grouped if they were reported in ten or more analyses using a compatible staffing measure. When less than twelve analyses reported on the same outcome, we grouped outcomes in common themes.

## RESULTS

Our search yielded 4,080 records, of which 954 were duplicates and 2,834 were excluded based on title and abstract. We assessed the full text of the remaining 292 studies for eligibility. Of these, 28 were included in the review (see Figure 1).

**Figure 1.** PRISMA Flow Diagram adapted from Page et al. The PRISMA 2020 statement: an updated guideline for reporting systematic reviews. BMJ 2021;372:n71. doi: 10.1136/bmj.n71, distributed under the terms of the Creative Commons Attribution License

Studies were published between 2003 and 2021. Most studies collected data from one hospital only (n= 15), but there were some notable exceptions, for example McHugh et al included 55 hospitals,^19^ Hamilton et al included 54 hospitals,^20^ and Mark and Belyea included 145 hospitals. Patient samples ranged between 85^14^ and 489,155.^19^ In terms of settings, 11 studies were conducted in Intensive Care Units (ICU); three in Acute Medical Units; thirteen in a variety of inpatient wards, which could also include ICUs. Most studies (n=9) were conducted in the US, 5 in the UK, 4 in Canada, 3 each in Italy, Australia and Switzerland, and one in Finland.

The length of the gap between the measurement of staffing levels and the outcome (the “exposure window”) ranged between 6 hours^21^ and 30 days.^15^ In studies considering policy implementations, staffing and outcomes relationships were measured one year after implementation.^18^ ^19^ Thirteen studies considered staffing levels averaged or summed cumulatively for the whole patient stay. ^13^ ^16^ ^17^ ^20^ ^22-30^ Six studies considered staffing in the early part of the patient stay only, ranging between the first day and the first seven days of the patients’ stay. ^23-25^ ^31-33^

Nurse staffing levels were measured in a variety of ways, which are summarised in Table 1 Staffing levels and skill mix measures deployed by studies. Some studies used multiple measures of staffing.

**Table 1.** Staffing levels and skill mix measures deployed by studies (table created by authors)

| Staffing measure | Number of studies |
| --- | --- |
| Nurse-to-patient ratio | 6 |
| Ratio of available to recommended nurses | 1 |
| Ratio of ICU trained to trainee nurses | 1 |
| Number of specialist nurses available divided by the number of recommended specialist nurses per shift | 1 |
| Number of nurses available divided by the number of recommended nurses per shift | 1 |
| Number of nurses per shift | 1 |
| Patient-to-nurse ratio | 1 |
| Nursing Hours per Patient Day (NHPPD) | 4 |
| RN HPPD relative to the ward mean | 3 |
| NA HPPD relative to the ward mean | 3 |
| NHPPD below 80% of the ward mean | 1 |
| NHPPD ≥ 8 hour below requirement | 1 |
| Nurse hours per patient per 12-h shift | 1 |
| Nurse hours per shift | 1 |
| Number of shifts with low nurse hours per patient per shift | 1 |
| Total number of RN hours worked divided by the total number of required hours of care based on patient dependency categories | 1 |
| Shifts with nurse hours lower than expected by ≥ 8 hours | 1 |
| Number of shifts with low nurse hours per patient shift | 1 |
| High / low staffing (50% above / below median number of RN expected given ward, patient activity & shift type ) | 1 |
| Number of nurses providing care to a patient prior to an event | 1 |
| Being cared for by float (temporary) nurses for % of the time | 1 |
| Days with additional Temporary NHPPD | 1 |
| Skill mix per shift (NonRN/RN+NonRN) | 1 |
| Skill mix (RNs/RNs+LPNs+NAs) | 1 |
| Skill mix (NA/RNs+LPN+NAs) | 1 |
| Skill mix (LPN/RNs+LPNs+NAs) | 1 |

RN: Registered Nurse; NA: Nursing Assistant; LPN: Licensed Practical Nurse; HPPD: Hours per Patient Day

### Risk of bias

Of the 28 studies, only three were assessed as at overall low risk of bias, seven studies were at moderate risk of bias, and twelve studies were classified as at serious risk of bias. Finally, six studies were at critical risk of bias and were not included in our synthesis.

Overall, we found that most studies had either serious (n=6) or critical (n=3) risk of bias resulting from confounding. I Many studies lacked appropriate adjustment for risk at baseline and/or failed to address time-varying confounding resulting from potential staffing increases as a response to worsening of a patients’ condition. Bias in selection of participants into the study was either low or moderate, apart from two studies of the implementation of staffing policies, which were classified as serious risk of bias due to the selection of units and hospitals which implemented staffing interventions and controls, which was not random. ^18^ ^19^

In many studies, the direction of any bias was determined to be unpredictable. However, in 9 studies with a serious or critical risk of bias, lack of individual risk-adjustment would likely bias results in favour of lower staffing or attenuate any observed effects from higher staffing. Failure to include known predictors of the outcome in statistical models could have led to estimates favouring lower staffing levels because patients at low risk of experiencing a negative outcome have lower need and may consequently be exposed to lower staffing. ^34^ ^35^ On the other hand, in two studies failure to control for levels of other staff could lead to an overestimate of the benefits of higher nurse staffing levels because the nurse staffing measure was likely to correlate with staffing by other staff groups and these staff groups have an effect on outcomes. While some other studies had no direct control for other staff groups the effect of the omission was less predictable as the design meant that variation in nurse staffing was unlikely to be correlated with that of other staff groups or else there was evidence presented for a lack of correlation. Detailed assessments can be found in Appendix 3.

### Mortality

Most studies and analyses showed higher staffing to be associated with reduced mortality. Twelve studies reported 40 analyses on patient mortality, but two studies (8 analyses) were at critical risk of bias and were excluded from the narrative synthesis.

Focusing on RNs (20 analyses in total), most studies and analyses showed higher staffing to be associated with reduced mortality although there was some inconsistency. Studies favouring higher staffing included larger studies at low/moderate risk of bias. Thirteen analyses from nine studies found that higher RN staffing levels were associated with a reduced risk of in-hospital mortality in adult general patient populations^13^ ^23-26^ ^28^ ^30^ ^32^ and reduced risk of death in neonatal populations.^20^ Six analyses from four studies reported associations between RN staffing levels and patient mortality that were not statistically significant, ^20^ ^25^ ^26^ ^33^ although coefficients, when presented, favoured higher RN staffing. One analysis from one small study with serious risk of bias found that higher RN staffing levels were associated with higher risk of mortality.^33^

We report all point estimates for RN staffing and mortality in Table *2*. Effect sizes were, typically, small. Exposure to low staffing was measured with different thresholds and different exposure windows. The diversity of staffing measures and exposure windows makes any meaningful comparison or synthesis impossible.

**Table 2.** Estimates for the effects of variation of RN staffing levels of mortality (table created by authors)

| Authors, year | Setting | Sample size | Staffing measure | Exposure window | Outcome | Result | Risk of Bias |
| --- | --- | --- | --- | --- | --- | --- | --- |
| Effect of lower RN staffing |  |  |  |  |  |  |  |
| Callaghan et al, 2003 | Neonatal Intensive Care Unit | **Hospitals** = 1; **Wards** = 1; **Patients**= 692 | Patient to nurse ratio of 1.71-1.97 | Averaged across first 72 hours of stay | In-hospital mortality | Compared to exposure to ratio 1.16-1.58: OR= 0.18; 95% CI= 0.06–0.5 * | Serious |
| Callaghan et al, 2003 | Neonatal Intensive Care Unit | **Hospitals** = 1; **Wards** = 1; **Patients**= 692 | Patient to nurse ratio of 1.59-1.70 | Averaged across first 72 hours of stay | In-hospital mortality | Compared to exposure to ratio 1.16-1.58: OR= 0.84; 95% CI= 0.42–1.66 | Serious |
| Griffiths et al, 2019 | Inpatient medical and surgical wards | **Hospitals** = 1; **Wards** = 32; **Patients** = 138,133; **RNs** = 1244; **NAs** = 700 | Days with staffing below the unit mean | Cumulative sum of days over the first 5 days of stay | In-hospital mortality | HR = 1.03; 95% CI = 1.01-1.06 * | Low |
| Needleman et al, 2011 | Intensive care, step-down centre, and general care | **Hospitals** = 1; **Wards** = 43; **Admissions** = 197,961; **Shifts** = 176,696; **Patient unit-shifts**= 3,227,457 | NHPPD ≥ 8 hour below requirement | Cumulative sum across all stay | In-hospital mortality | HR = 1.02; 95% CI = 1.01 - 1.03 * | Moderate |
| Needleman et al, 2011 | Intensive care, step-down centre, and general care | **Hospitals** = 1; **Wards** = 43; **Admissions** = 197,961; **Shifts** = 176,696; **Patient unit-shifts**= 3,227,457 | NHPPD ≥ 8 hour below requirement | Cumulative sum of first five days of patient stay | In-hospital mortality | HR = 1.03; 95% CI = 1.02 - 1.05 * | Moderate |
| Needleman et al, 2011 | Intensive care, step-down centre, and general care | **Hospitals** = 1; **Wards** = 43; **Admissions** = 197,961; **Shifts** = 176,696; **Patient unit-shifts**= 3,227,457 | NHPPD ≥ 8 hour below requirement | Cumulative sum of 2 shifts prior to death | In-hospital mortality | HR = 1.05; 95% CI = 1.02 - 1.07* | Moderate |
| Needleman et al, 2020 | Adult, nonsurgical and non-psychiatric | **Hospitals**= 3 (2 tertiary and 1 community); **Admissions**= 78,303 | Nurse hours per patient per 12-h shift | Any shift with staffing below 75% of annual median unit | In-hospital mortality | HR = 1.02; 95% CI = 1.00 -1.05* | Moderate |
| Needleman et al, 2020 | Adult, nonsurgical and non-psychiatric | **Hospitals**= 3 (2 tertiary and 1 community);<br>**Admissions**= 78,303 | Number of shifts with low nurse hours per patient per shift | Cumulative sum over the 2nd to 5th day of stay | In-hospital mortality | HR = 1.04; 95% CI = 0.99; 1.10 | Moderate |
| Rochefort et al, 2020 | Adult medical, surgical, and intensive care units | **Hospitals**= 1; **Wards**= 32;<br>**Patients**= 146,349 | Shifts with nurse hours lower than expected by $\geq 8$ hours | Cumulative sum across stay | In-hospital mortality | HR = 1.01; 95% CI = 1.00 -1.01* | Low |
| **Effect of higher RN staffing** |  |  |  |  |  |  |  |
| Beltempo et al, 2018 | Neonatal Intensive Care Unit | **Hospitals**= 1; **Wards**= 1; **Nurses**= 165;<br>**patients**= 257 | Total number of nurse hours worked divided by the total number of required hours of care based on patient dependency categories | First day of NICU stay | Mortality (all causes) or major morbidity | RR= 0.83; 95% CI= 0.77–0.90* | Moderate |
| Beltempo et al, 2018 | Neonatal Intensive Care Unit | **Hospitals**= 1; **Wards**= 1; **Nurses**= 165;<br>**patients**= 257 | Number of RN hours worked divided by the number of required hours of care based on patient dependency categories | Averaged across first 7 days | Mortality (all causes) or major morbidity | RR= 0.82; 95% CI: 0.76–0.89* | Moderate |
| Beltempo et al, 2018 | Neonatal Intensive Care Unit | **Hospitals**= 1; **Wards**= 1; **Nurses**= 165;<br>**patients**= 257 | Total number of nurse hours worked divided by the total number of required hours of care based on patient dependency categories | Averaged across patient stay | Mortality (all causes) or major morbidity | RR= 0.81; 95% CI= 0.74-0.90* | Moderate |
| Fogg et al, 2021 | Inpatient medical and surgical wards, patients >75 yrs old | **Hospitals** = 1; **Wards** = 32; **Patients** = 9,643 | RN hours per patient day relative to the mean (with non-linear terms) | Averaged across patient stay relative to the mean | In-hospital mortality and within 30 days of discharge | Mean vs mean + 0.5 RNHPPD: OR= 0.90; 95% CI = 0.84–0.97* | Moderate |
| Griffiths et al, 2019 | Inpatient medical and surgical wards | **Hospitals** = 1; **Wards** = 32; **Patients** = 138,133; **RNs** = 1244; **NAs** = 700 | RN hours per patient day relative to the mean | Cumulative sum over first 5 days of stay | In-hospital mortality | HR = 0.97; 95% CI = 0.94 –0.99* | Low |
| Hamilton et al, 2007 | Neonatal Intensive Care Unit | **Hospitals**= 54;<br>**Wards**= 54;<br>**Patients**= 2585 | Number of specialist nurses available divided by the number of recommended specialist nurses per shift | Averaged across patient stay | Death before discharge and planned deaths at home | OR = 0.63; 95% CI = 0.42 - 0.96* | Moderate |
| Musy et al, 2021 | Adult inpatients | **Hospital** = 1; **Wards** =55; **Patients** = 79,893; **RNs** = 3646; **LPNs** = 438 | High / low staffing (50% above / below median number of RN expected given ward, patient activity & shift type) | Cumulative count of shifts with high numbers across all stay | In-hospital Mortality | Higher staffing: OR = 0.91*; 95% CI = 0.89-0.93 | Low |
| Hamilton et al, 2007 | Neonatal Intensive Care Unit | **Hospitals**= 54; **Wards**= 54; **Patients**= 2585 | Number of nurses per shift | Averaged across patient stay | Death before discharge and planned deaths at home | No estimates - reported as non significant | Moderate |
| Hamilton et al, 2007 | Neonatal Intensive Care Unit | **Hospitals**= 54; **Wards**= 54; **Patients**= 2585 | Number of nurses available divided by the number of recommended nurses per shift | Averaged across patient stay | Death before discharge and planned deaths at home | No estimates - reported as non significant | Moderate |
| Rocheftort et al, 2020 | Adult medical, surgical, and intensive care units | **Hospitals**= 1; **Wards**= 32; **Patients**= 146,349 | Nurse/patient ratio | Cumulative sum across stay | In-hospital mortality | No estimates - reported as non significant | Low |
| Rocheftort et al, 2020 | Adult medical, surgical, and intensive care units | **Hospitals**= 1; **Wards**= 32; **Patients**= 146,349 | Nurse hours per shift | Cumulative sum across stay | In-hospital mortality | No estimates - reported as non significant | Low |

\* = statistically significant at <0.05; OR = Odds Ratio; HR = Hazard Ratio; RR = Risk Ratio; CI = Confidence Interval

Skill mix and non-RN staffing findings were mixed and provide an inconsistent picture with contrasting results. Three analyses (two studies) reported increased mortality when patients were exposed to low NA staffing, ^25^ ^32^ although Griffiths et al found a non-linear relationship so NA staffing above the norm was also associated with increased risk of mortality.^32^ Two analyses from two studies reported higher mortality when patients were exposed to higher NA staffing levels, although these were not statistically significant.^28^ ^32^ Needleman and colleagues also combined RN and NA staffing levels in two analyses and found patients exposed to lower staffing levels from both groups were more likely to die.^25^ Musy et al found that being exposed to either high or low LPN staffing levels was associated with lower mortality, although only high LPN staffing levels were statistically significant.^30^ Skill mix was not associated with patient mortality.^26^ Point estimates for all nurses, NA and LPN staffing and mortality analyses are reported in Table 3.

**Table 3.**
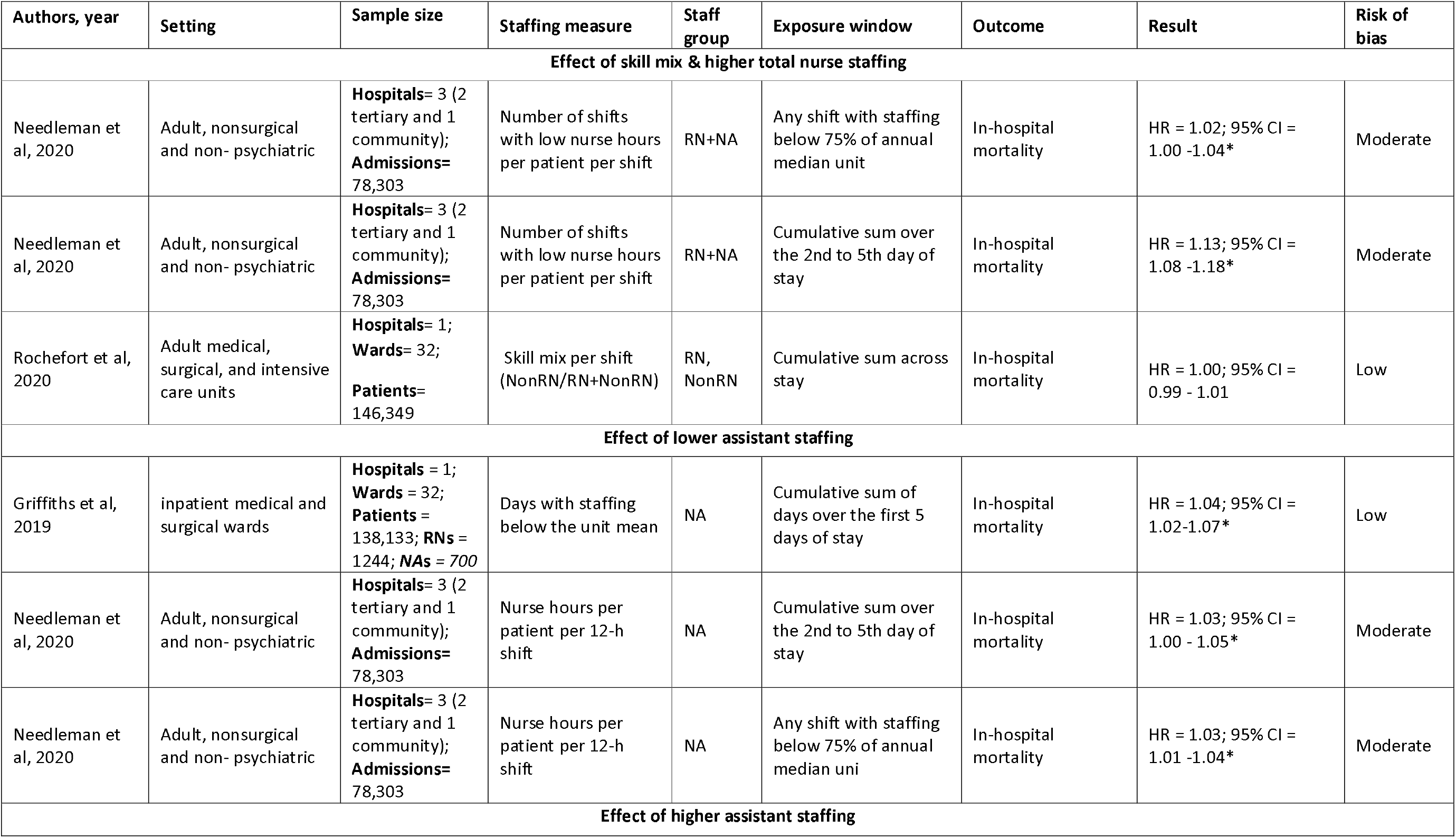
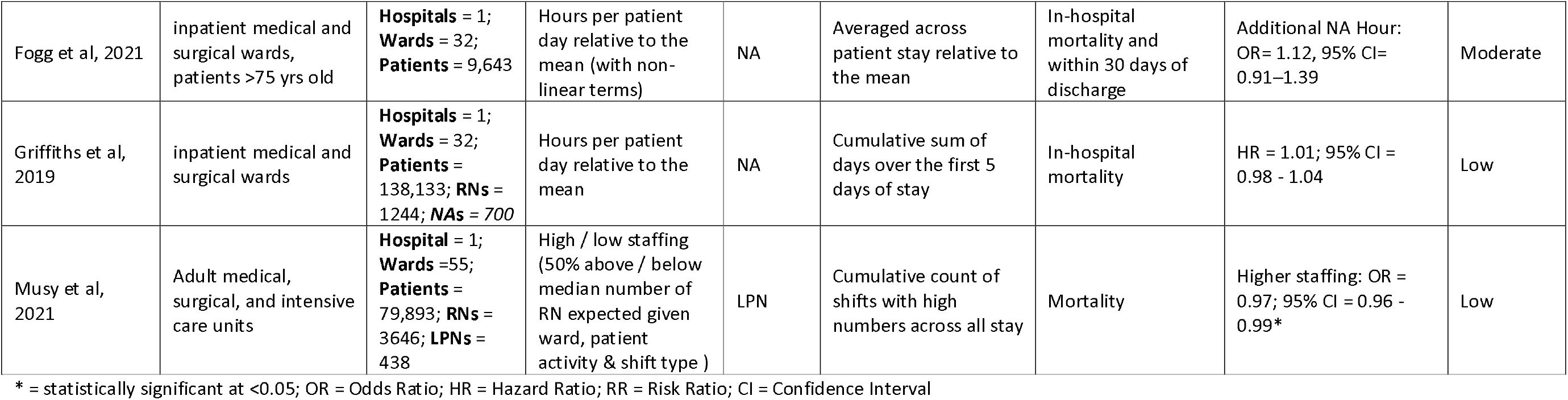
Point estimates for all nurses, NA, LPN staffing and mortality (table created by authors)

Focusing on temporary staffing, three analyses from one study in one hospital with 138,133 found that exposure to higher levels of temporary RN and NA staffing were associated with higher patient mortality. However, for RNs this effect was found only when a high number of RN hours, equivalent to between one third and one half of the average ward staffing complement, were from temporary nurses (HR = 1.12; 95% CI = 1.03–1.21).^31^ The study was at serious risk of bias.

### Infections

Overall, there was inconclusive evidence of the effect of nurse staffing levels on infections, with results pointing towards an effect in favour of higher RN staffing levels.

After removing one study at critical risk of bias, 16 analyses from seven studies examined the impact of staffing levels on patients’ infections, including central venous catheter (CVC)-associated bloodstream infections, early and late onset ventilator-associated pneumonia, and healthcare-associated infections. Considering RNs, eight analyses from seven studies reported statistically non-significant results, although point estimates, when available, showed a protective effect of higher RN staffing,^22^ ^36-39^ except for one analysis.^40^ ^41^ Four analyses found that being exposed to higher RN staffing was protective of infections in neonatal intensive care settings,^40^ ^41^ adult intensive care settings^37^ ^38^ and adult hospital settings.^39^

While two analyses found no statistically significant relationships (point estimates not reported) between NA staffing and infections,^22^ ^39^ one analysis found that higher NA staffing levels were associated with lower risk of healthcare-associated infections.^39^ One analysis found that patients exposed to days with higher levels of temporary nurse staffing were more likely to experience CVC-associated bloodstream infections.^22^ All available point estimates for the analyses of staffing levels and infections are reported in Table 4.

**Table 4.**
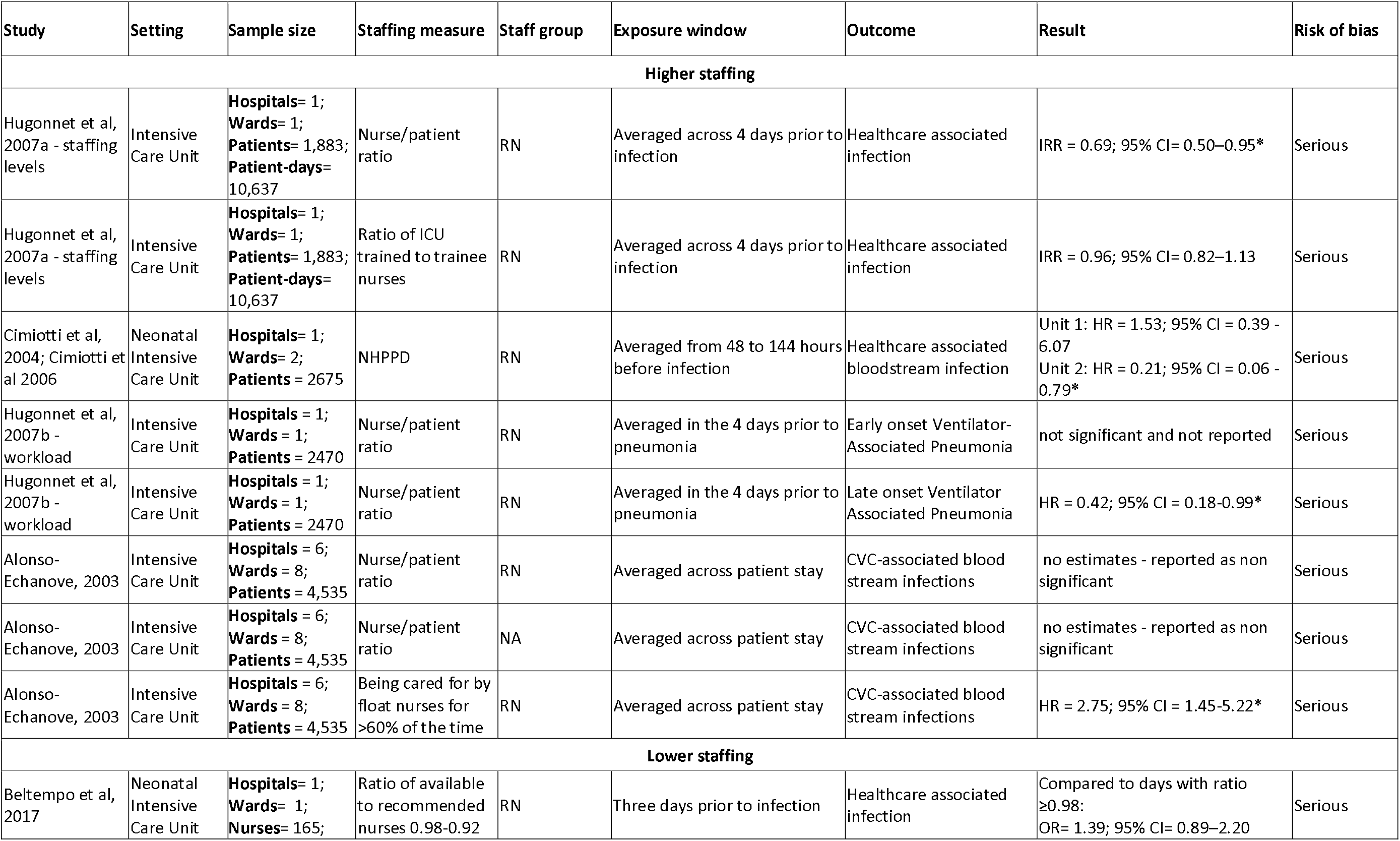
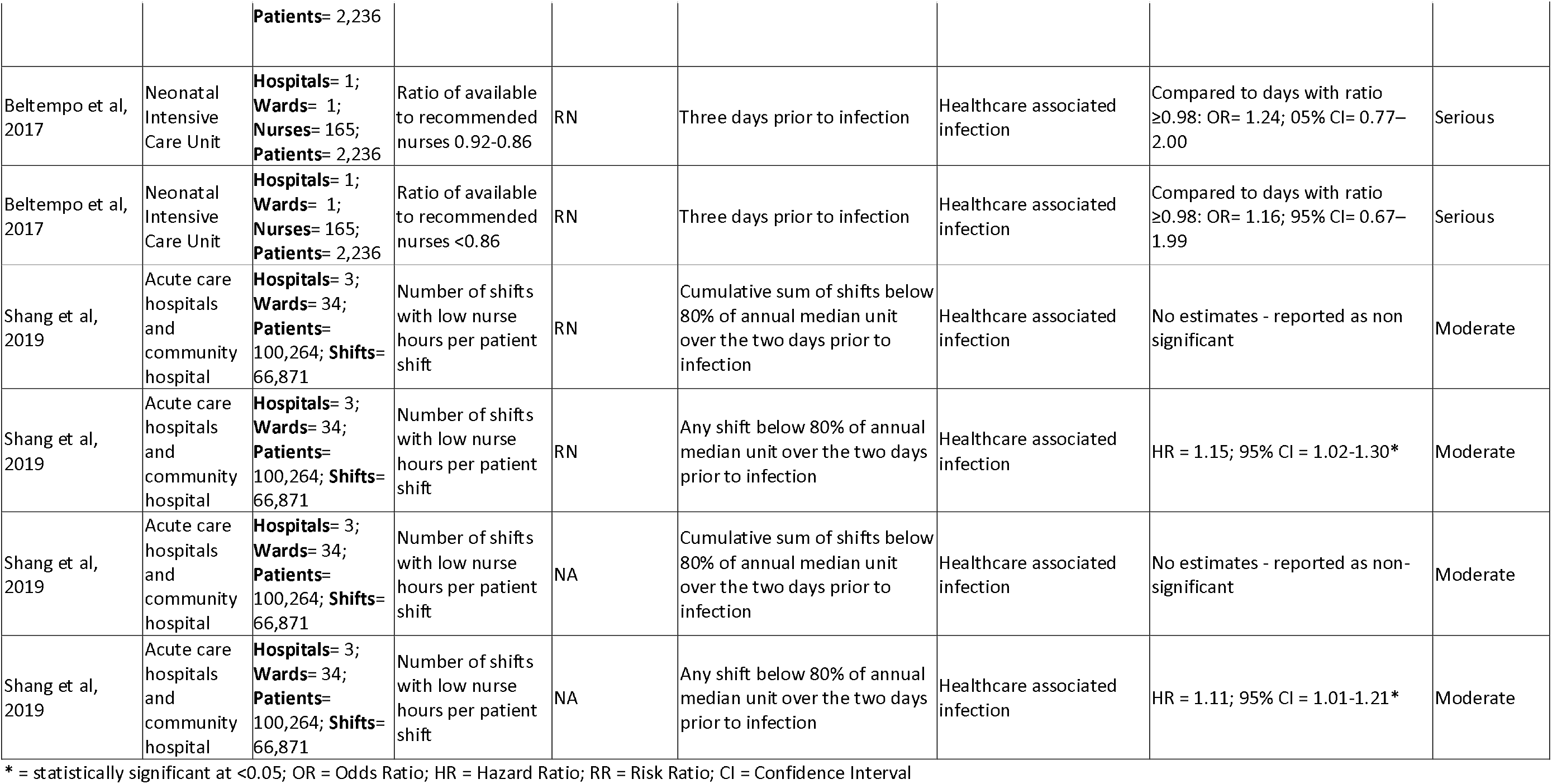
Point estimates for analyses of staffing levels and infections (table created by authors)

| Study | Setting | Sample size | Staffing measure | Staff group | Exposure window | Outcome | Result | Risk of bias |
| --- | --- | --- | --- | --- | --- | --- | --- | --- |
| **Higher staffing** |  |  |  |  |  |  |  |  |
| Hugonnet et al, 2007a - staffing levels | Intensive Care Unit | **Hospitals= 1;**<br>**Wards= 1;**<br>**Patients= 1,883;**<br>**Patient-days= 10,637** | Nurse/patient ratio | RN | Averaged across 4 days prior to infection | Healthcare associated infection | IRR = 0.69; 95% CI= 0.50–0.95* | Serious |
| Hugonnet et al, 2007a - staffing levels | Intensive Care Unit | **Hospitals= 1;**<br>**Wards= 1;**<br>**Patients= 1,883;**<br>**Patient-days= 10,637** | Ratio of ICU trained to trainee nurses | RN | Averaged across 4 days prior to infection | Healthcare associated infection | IRR = 0.96; 95% CI= 0.82–1.13 | Serious |
| Cimiotti et al, 2004; Cimiotti et al 2006 | Neonatal Intensive Care Unit | **Hospitals= 1;**<br>**Wards= 2;**<br>**Patients = 2675** | NHPPD | RN | Averaged from 48 to 144 hours before infection | Healthcare associated bloodstream infection | Unit 1: HR = 1.53; 95% CI = 0.39 - 6.07<br>Unit 2: HR = 0.21; 95% CI = 0.06 - 0.79* | Serious |
| Hugonnet et al, 2007b - workload | Intensive Care Unit | **Hospitals = 1;**<br>**Wards = 1;**<br>**Patients = 2470** | Nurse/patient ratio | RN | Averaged in the 4 days prior to pneumonia | Early onset Ventilator-Associated Pneumonia | not significant and not reported | Serious |
| Hugonnet et al, 2007b - workload | Intensive Care Unit | **Hospitals = 1;**<br>**Wards = 1;**<br>**Patients = 2470** | Nurse/patient ratio | RN | Averaged in the 4 days prior to pneumonia | Late onset Ventilator Associated Pneumonia | HR = 0.42; 95% CI = 0.18-0.99* | Serious |
| Alonso-Echanove, 2003 | Intensive Care Unit | **Hospitals = 6;**<br>**Wards = 8;**<br>**Patients = 4,535** | Nurse/patient ratio | RN | Averaged across patient stay | CVC-associated blood stream infections | no estimates - reported as non significant | Serious |
| Alonso-Echanove, 2003 | Intensive Care Unit | **Hospitals = 6;**<br>**Wards = 8;**<br>**Patients = 4,535** | Nurse/patient ratio | NA | Averaged across patient stay | CVC-associated blood stream infections | no estimates - reported as non significant | Serious |
| Alonso-Echanove, 2003 | Intensive Care Unit | **Hospitals = 6;**<br>**Wards = 8;**<br>**Patients = 4,535** | Being cared for by float nurses for >60% of the time | RN | Averaged across patient stay | CVC-associated blood stream infections | HR = 2.75; 95% CI = 1.45-5.22* | Serious |
| **Lower staffing** |  |  |  |  |  |  |  |  |
| Beltempo et al, 2017 | Neonatal Intensive Care Unit | **Hospitals= 1;**<br>**Wards= 1;**<br>**Nurses= 165;** | Ratio of available to recommended nurses 0.98-0.92 | RN | Three days prior to infection | Healthcare associated infection | Compared to days with ratio ≥0.98:<br>OR= 1.39; 95% CI= 0.89–2.20 | Serious |
|  |  | **Patients= 2,236** |  |  |  |  |  |  |
| Beltempo et al, 2017 | Neonatal Intensive Care Unit | **Hospitals= 1; Wards= 1; Nurses= 165; Patients= 2,236** | Ratio of available to recommended nurses 0.92-0.86 | RN | Three days prior to infection | Healthcare associated infection | Compared to days with ratio $\geq 0.98$: OR= 1.24; 05% CI= 0.77–2.00 | Serious |
| Beltempo et al, 2017 | Neonatal Intensive Care Unit | **Hospitals= 1; Wards= 1; Nurses= 165; Patients= 2,236** | Ratio of available to recommended nurses <0.86 | RN | Three days prior to infection | Healthcare associated infection | Compared to days with ratio $\geq 0.98$: OR= 1.16; 95% CI= 0.67–1.99 | Serious |
| Shang et al, 2019 | Acute care hospitals and community hospital | **Hospitals= 3; Wards= 34; Patients= 100,264; Shifts= 66,871** | Number of shifts with low nurse hours per patient shift | RN | Cumulative sum of shifts below 80% of annual median unit over the two days prior to infection | Healthcare associated infection | No estimates - reported as non significant | Moderate |
| Shang et al, 2019 | Acute care hospitals and community hospital | **Hospitals= 3; Wards= 34; Patients= 100,264; Shifts= 66,871** | Number of shifts with low nurse hours per patient shift | RN | Any shift below 80% of annual median unit over the two days prior to infection | Healthcare associated infection | HR = 1.15; 95% CI = 1.02-1.30* | Moderate |
| Shang et al, 2019 | Acute care hospitals and community hospital | **Hospitals= 3; Wards= 34; Patients= 100,264; Shifts= 66,871** | Number of shifts with low nurse hours per patient shift | NA | Cumulative sum of shifts below 80% of annual median unit over the two days prior to infection | Healthcare associated infection | No estimates - reported as non-significant | Moderate |
| Shang et al, 2019 | Acute care hospitals and community hospital | **Hospitals= 3; Wards= 34; Patients= 100,264; Shifts= 66,871** | Number of shifts with low nurse hours per patient shift | NA | Any shift below 80% of annual median unit over the two days prior to infection | Healthcare associated infection | HR = 1.11; 95% CI = 1.01-1.21* | Moderate |

\* = statistically significant at <0.05; OR = Odds Ratio; HR = Hazard Ratio; RR = Risk Ratio; CI = Confidence Interval

### Other outcomes

In a single site study of 138,133 patients, Griffiths and colleagues examined the effect of staffing levels on a composite outcome of adverse events, including death, cardiac arrest or unplanned ICU admission. They found that being exposed to higher RNHPPD in the first five days of a hospital admission was associated with a reduced hazard of experiencing adverse events (HR = 0.98; 95% CI = 0.96-0.99). Results for NAHPPD were in the opposite direction, although not statistically significant (HR = 1.01; 95% CI = 0.99-1.02).^29^

Patrician and colleagues examined the effect of staffing levels and skill mix on hospital-acquired pressure injuries in 13 hospitals and 1643 patients through 12 distinct analyses, exploring various exposure windows and skill mix configurations. The majority (10/12) of these analyses found no statistically significant associations, but two analyses found that a skill mix richer in LPNs over the three days (HR = 0.27 (no 95% CI reported)) and one week prior (HR = 0.56 (no 95% CI reported)) to the hospital-acquired pressure injury was protective of hospital-acquired pressure injuries.^42^

One study in two ICUs (one paediatric and one cardiac) and 11,310 admissions found that patients were more likely to receive positive pressure ventilation following unplanned extubation when there were more RNs in the preceding six hours (OR= 1.53; 95% CI: 1.11-2.12).^21^

Griffiths and colleagues found that patients’ length of stay in hospital was reduced by a mean of 0.23 days for each additional RN HPPD that a patient experienced (Gamma coefficient = -0.23; 95% CI = - 0.30 - -0.16), while there was a minimal increase in days for each additional NA HPPD a patient was exposed to throughout their stay (Gamma coefficient = 0.076; 95% CI = 0.03 - 0.13).^29^ Tschannen found that being exposed to higher NHPPD was associated with shorter than expected stays (relative to Diagnoses Related Groups based norms) (B= 2.481, SE= 1.0), but not overall average length of stay (B =0.43, SE = 0.01).^27^ A single site study of 9,643 patients over the age of 75 who received a cognitive screening, found that, for each day patients were exposed to an additional 0.5 RNHPPD, they were less likely to be readmitted, although this was not statistically significant (OR = 0.94; 95% CI = 0.82–1.06).^28^

### Staffing policies

Of two studies examining the effect of staffing policy interventions,^18^ ^19^ one was at critical risk of bias. A prospective panel study compared patient outcomes in hospitals that implemented a minimum nurse-to-patient ratio policy. Twenty-seven hospitals were subject to the staffing policy and 28 were not. Reducing workloads by one patient per nurse was associated with a decrease in 30-day mortality (OR= 0.93; 95% CI = 0.86-0.99), 7-day readmissions (OR = 0.93; 95% CI = 0.89-0.97) and length of stay (OR= 0.97; 95% CI = 0.94-0.99).^19^

## DISCUSSION

This is the first systematic review of the effect of nurse staffing levels on patient outcomes focusing on longitudinal studies that can demonstrate a temporal link between exposure to staffing levels and outcomes. While studies were performed in different populations and some were at serious or critical risk of bias, findings are consistent with higher RN staffing reducing the risk of patient death. On the other hand, evidence about the positive contribution of other staff groups and the skill mix of the team is unclear.

This evidence is consistent with conclusions based primarily on cross-sectional evidence;^3^ but our review is limited to studies that can demonstrate that low staffing levels precede outcomes and thus eliminate several sources of confounding. When considering criteria to establish a causal relationship, there is a fundamental requirement that cause precedes effect.^43^ ^44^ Although the evidence reviewed here remains observational, if relevant confounders are controlled for in the analyses, the parameter estimates for exposure can be causally interpreted,^45^ although some risk of bias remains. Studies with the greatest risk of bias were judged as most likely to underestimate the effect of higher nurse staffing.

The diverse measures of exposure and diverse settings and patient populations make it difficult to come to firm conclusions about the size of effects beyond the estimates provided by individual studies. However, while the effect sizes observed for mortality are typically small, the large populations exposed to risk means that these effects are, nonetheless, important. In a study that included 138,000 general medical and surgical admissions to one large hospital over 3 years we estimated that an increase of 1 RN hour per patient day might avoid 657 deaths and reduce bed use by over 30,000 bed days.^29^

When considering NAs and other grades of nursing staff, the effects observed are mixed, and so a causal inference from these results is more challenging. It seems likely from the evidence that any causal relationship is complex, with the hints of non-linear effects^32^ suggesting competing causal mechanisms from increased resource as opposed to substitution of RNs. Findings around temporary staffing, albeit from a single study, show that when small proportions of temporary nurses are deployed, possibly to maintain staffing levels, the effect on mortality seems to be beneficial, but when between a third and half of the RNs on shift are temporary, the association with mortality changes direction.

Regarding other patient outcomes, there is more uncertainty due to higher risk of bias in studies; while the direction of the relationships observed remains compatible with a protective effect of higher nurse staffing levels, most studies are at either serious or critical risk of bias.

### Recommendations for future research practice

The evidence for the link between nurse staffing levels and mortality is strong from an internal validity perspective, but longitudinal studies using routinely collected data from a larger number of hospitals are needed to increase external validity, improving both the precision and generalisability of estimates. While we did not formally assess external validity, all studies at low risk of bias were conducted in a single hospital, which limits generalisability of findings. Ensuring external validity will be crucial to change policy and practice^46^ ^47^ around nurse staffing levels.

Considering other patient outcomes, studies that can address the limitations of existing literature are needed. The causal pathway from low staffing to mortality has been theorised and empirically demonstrated, in part mediated by a failure to observe and mobilise response to deterioration.^48-51^ While much work has been done already about nurse-sensitive outcomes,^52-54^ the current evidence does not provide a consistent and coherent overview of how nurse staffing affects other patient outcomes. Estimation of the effect of variation in nurse staffing levels on some well-established nurse sensitive outcomes including falls and pressure ulcers is hampered by inadequate risk adjustment models, poor recording, and ascertainment bias in the current research.

In addition, we note that staffing levels were measured in a variety of ways across studies. If approaches to measuring staffing inputs were standardised, or if raw anonymised data were provided alongside papers, more comparable estimates could be combined in meta-analyses. While it is difficult to establish the clear superiority of one staffing effect measure over another, we would encourage all authors to offer analyses using measures based on existing reports in addition to any de-novo measures they derive. Staffing deviation standardised against unit means has been used as an absolute (low staffing) and continuous (hours per patient day relative to the mean) effect measure, which could be derived for all studies irrespective of the methods used to determine staffing requirements.

### Limitations of the review

Although our search was extensive, the topic is difficult to capture precisely in structured searches. Although as a team we are familiar with the literature and were thus able to test search strategies, it is possible that we missed some eligible studies. However, it seems unlikely that we would have missed a sufficient number of low risk of bias studies to fundamentally change our conclusions or the general picture of the literature that we have presented here.

## CONCLUSIONS

This review identified a likely causal relationship between higher RN staffing levels and reduced patient death. In contrast, we found much less certain evidence about the role of assistant staff, changes in skill mix and grades of nurse other than RNs. While it seems possible that such staff may make some contribution to patient safety, the evidence supports a policy of increases in RNs in the acute setting studied but cannot be used to support substitution of RNs by other grades of staff. That there is a causal relationship between low RN staffing and harm to patients now seems beyond dispute although uncertainties remain regarding the magnitude of effect.

## Data Availability

No data included - systematic review

## REFERENCES

1. World Health Organization. Global strategic directions for strengthening nursing and midwifery 2016-2020. 2016

2. Yakusheva O, Rambur B, Buerhaus PI. Value-Informed Nursing Practice Can Help Reset the Hospital-Nurse Relationship. JAMA Health Forum 2020;1(8):e200931–e31. doi: 10.1001/jamahealthforum.2020.0931

3. Griffiths P, Ball J, Drennan J, et al. Nurse staffing and patient outcomes: Strengths and limitations of the evidence to inform policy and practice. A review and discussion paper based on evidence reviewed for the National Institute for Health and Care Excellence Safe Staffing guideline development. Int J Nurs Stud 2016;63:213–25. doi: 10.1016/j.ijnurstu.2016.03.012 [published Online First: 2016/05/01]

4. Buchan J, Ball J, Shembavnekar N, et al. Building the NHS nursing workforce in England. 2020 doi: https://doi.org/10.37829/HF-2020-RC14

5. Beech J, Bottery S, Charlesworth A, et al. Closing the gap. Key areas for action on the health and care workforce London: The Health Foundation/Nuffield Trust/The King’s Fund 2019

6. Kane RL, Shamliyan TA, Mueller C, et al. The association of registered nurse staffing levels and patient outcomes: systematic review and meta-analysis. Med Care 2007;45(12):1195–204. doi: 10.1097/MLR.0b013e3181468ca3 [published Online First: 2007/11/17]

7. Shekelle PG. Nurse–patient ratios as a patient safety strategy: a systematic review. Ann Intern Med 2013;158(5_Part_2):404–09.

8. Driscoll A, Grant MJ, Carroll D, et al. The effect of nurse-to-patient ratios on nurse-sensitive patient outcomes in acute specialist units: a systematic review and meta-analysis. Eur J Cardiovasc Nurs 2018;17(1):6–22.

9. Shin S, Park JH, Bae SH. Nurse staffing and hospital-acquired conditions: A systematic review. J Clin Nurs 2019;28(23-24):4264–75.

10. Musy SN, Endrich O, Leichtle AB, et al. Longitudinal Study of the Variation in Patient Turnover and Patient-to-Nurse Ratio: Descriptive Analysis of a Swiss University Hospital. Journal of Medical Internet Research 2020;22(4):N.PAG-N.PAG. doi: 10.2196/15554

11. Haegdorens F, Van Bogaert P, De Meester K, et al. The impact of nurse staffing levels and nurse’s education on patient mortality in medical and surgical wards: an observational multicentre study. BMC Health Serv Res 2019;19(1):864. doi: 10.1186/s12913-019-4688-7 [published Online First: 2019/11/23]

12. Sterne JA, Hernán MA, Reeves BC, et al. ROBINS-I: a tool for assessing risk of bias in non-randomised studies of interventions. BMJ (Clinical research ed) 2016;355:i4919–i19. doi: 10.1136/bmj.i4919

13. Ambrosi E, De Togni S, Guarnier A, et al. In-hospital elderly mortality and associated factors in 12 Italian acute medical units: findings from an exploratory longitudinal study. Aging Clin Exp Res 2017;29(3):517–27. doi: 10.1007/s40520-016-0576-8 [published Online First: 2016/05/09]

14. Jansson MM, Syrjälä HP, Ala-Kokko TI. Association of nurse staffing and nursing workload with ventilator-associated pneumonia and mortality: a prospective, single-center cohort study. J Hosp Infect 2019;101(3):257–63. doi: 10.1016/j.jhin.2018.12.001

15. Mark BA, Belyea M. Nurse staffing and medication errors: cross-sectional or longitudinal relationships? Res Nurs Health 2009;32(1):18–30. doi: 10.1002/nur.20305

16. Palese A, Ambrosi E, Prosperi L, et al. Missed nursing care and predicting factors in the Italian medical care setting. Intern Emerg Med 2015;10(6):693–702. doi: 10.1007/s11739-015-1232-6 [published Online First: 2015/04/05]

17. Palese A, Gonella S, Moreale R, et al. Hospital-acquired functional decline in older patients cared for in acute medical wards and predictors: Findings from a multicentre longitudinal study. Geriatr Nurs 2016;37(3):192–9. doi: 10.1016/j.gerinurse.2016.01.001 [published Online First: 2016/02/21]

18. Twigg DE, Myers H, Duffield C, et al. The impact of adding assistants in nursing to acute care hospital ward nurse staffing on adverse patient outcomes: An analysis of administrative health data. Int J Nurs Stud 2016;63:189–200. doi: 10.1016/j.ijnurstu.2016.09.008 [published Online First: 2016/09/23]

19. McHugh MD, Aiken LH, Sloane DM, et al. Effects of nurse-to-patient ratio legislation on nurse staffing and patient mortality, readmissions, and length of stay: a prospective study in a panel of hospitals. The Lancet 2021 doi: 10.1016/S0140-6736(21)00768-6

20. Hamilton KE, Redshaw MES, Tarnow-Mordi W. Nurse staffing in relation to risk-adjusted mortality in neonatal care. Archives of Disease in Childhood -- Fetal & Neonatal Edition 2007;92(2):F99–F103.

21. Al-Abdwani R, Williams CB, Dunn C, et al. Incidence, outcomes and outcome prediction of unplanned extubation in critically ill children: An 11year experience. J Crit Care 2018;44:368–75. doi: 10.1016/j.jcrc.2017.12.017 [published Online First: 2018/01/01]

22. Alonso-Echanove J, Edwards JR, Richards MJ, et al. Effect of nurse staffing and antimicrobial-impregnated central venous catheters on the risk for bloodstream infections in intensive care units. Infect Control Hosp Epidemiol 2003;24(12):916–25. doi: 10.1086/502160 [published Online First: 2004/01/01]

23. Beltempo M, Lacroix G, Cabot M, et al. Association of nursing overtime, nurse staffing and unit occupancy with medical incidents and outcomes of very preterm infants. J Perinatol 2018;38(2):175–80. doi: 10.1038/jp.2017.146

24. Needleman J, Buerhaus P, Pankratz VS, et al. Nurse staffing and inpatient hospital mortality. N Engl J Med 2011;364(11):1037–45. doi: 10.1056/NEJMsa1001025 [published Online First: 2011/03/18]

25. Needleman J, Jianfang L, Jinjing S, et al. Association of registered nurse and nursing support staffing with inpatient hospital mortality. BMJ Quality & Safety 2020;29(1):10–18. doi: 10.1136/bmjqs-2018-009219

26. Rochefort CM, Beauchamp M-E, Audet L-A, et al. Associations of 4 Nurse Staffing Practices With Hospital Mortality. Med Care 2020;58(10):912–18. doi: 10.1097/mlr.0000000000001397

27. Tschannen DJ. Organizational structure, process, and outcome: the effects of nurse staffing and nurse-physician collaboration on patient length of stay. University of Michigan, 2005.

28. Fogg C, Bridges J, Meredith P, et al. The association between ward staffing levels, mortality and hospital readmission in older hospitalised adults, according to presence of cognitive impairment: a retrospective cohort study. Age & Ageing 2021;50(2):431–39. doi: 10.1093/ageing/afaa133

29. Griffiths P, Ball J, Bloor K, et al. Nurse staffing levels, missed vital signs and mortality in hospitals: retrospective longitudinal observational study: NIHR Journals Library, Southampton (UK), 2018.

30. Musy SN, Endrich O, Leichtle AB, et al. The association between nurse staffing and inpatient mortality: a shift-level retrospective longitudinal study. International Journal of Nursing Studies 2021:103950. doi: https://doi.org/10.1016/j.ijnurstu.2021.103950

31. Dall’Ora C, Maruotti A, Griffiths P. Temporary Staffing and Patient Death in Acute Care Hospitals: A Retrospective Longitudinal Study. Journal of Nursing Scholarship 2020;52(2):210–16. doi: 10.1111/jnu.12537

32. Griffiths P, Maruotti A, Recio Saucedo A, et al. Nurse staffing, nursing assistants and hospital mortality: retrospective longitudinal cohort study. BMJ Quality & Safety 2019;28(8):609–17. doi: 10.1136/bmjqs-2018-008043

33. Callaghan LA, Cartwright DW, O’Rourke P, et al. Infant to staff ratios and risk of mortality in very low birthweight infants. Arch Dis Child Fetal Neonatal Ed 2003;88(2):F94–7. doi: 10.1136/fn.88.2.f94 [published Online First: 2003/02/25]

34. He J, Almenoff PL, Keighley J, et al. Impact of patient-level risk adjustment on the findings about nurse staffing and 30-day mortality in veterans affairs acute care hospitals. Nurs Res 2013;62(4):226–32. doi: 10.1097/NNR.0b013e318295810c [published Online First: 2013/07/03]

35. Antonakis J, Bendahan S, Jacquart P, et al. On making causal claims: A review and recommendations. Leadership Quarterly 2010;21(6):1086–120. doi: 10.1016/j.leaqua.2010.10.010

36. Beltempo M, Blais R, Lacroix G, et al. Association of Nursing Overtime, Nurse Staffing, and Unit Occupancy with Health Care-Associated Infections in the NICU. Am J Perinatol 2017;34(10):996–1002. doi: 10.1055/s-0037-1601459

37. Hugonnet S, Chevrolet J, Pittet D. The effect of workload on infection risk in critically ill patients. Crit Care Med 2007;35(1):76–81.

38. Hugonnet S, Uckay I, Pittet D. Staffing level: a determinant of late-onset ventilator-associated pneumonia. Crit Care 2007;11(4):R80. doi: 10.1186/cc5974 [published Online First: 2007/07/21]

39. Shang J, Needleman J, Liu J, et al. Nurse Staffing and Healthcare-Associated Infection, Unit-Level Analysis. J Nurs Adm 2019;49(5):260–65. doi: 10.1097/NNA.0000000000000748

40. Cimiotti JP. Nurse staffing and healthcare-associated infections in the neonatal ICU. Columbia University, 2004.

41. Cimiotti JP, Haas J, Saiman L, et al. Impact of staffing on bloodstream infections in the neonatal intensive care unit. Arch Pediatr Adolesc Med 2006;160(8):832–36. doi: DOI 10.1001/archpedi.160.8.832

42. Patrician PA, McCarthy MS, Swiger P, et al. Association of Temporal Variations in Staffing With Hospital-Acquired Pressure Injury in Military Hospitals. Res Nurs Health 2017;40(2):111–19. doi: 10.1002/nur.21781 [published Online First: 2016/12/10]

43. Hill AB. The Environment and Disease: Association or Causation? Proc R Soc Med 1965;58(5):295–300. [published Online First: 1965/05/01]

44. Rothman KJ, Greenland S. Causation and causal inference in epidemiology. Am J Public Health 2005;95 Suppl 1:S144–50. doi: 10.2105/ajph.2004.059204 [published Online First: 2005/07/21]

45. Hernán MA. The C-Word: Scientific Euphemisms Do Not Improve Causal Inference From Observational Data. American Journal of Public Health 2018;108(5):616–19. doi: 10.2105/ajph.2018.304337

46. Glasgow RE, Green LW, Klesges LM, et al. External validity: We need to do more. Ann Behav Med 2006;31(2):105–08. doi: 10.1207/s15324796abm3102_1

47. Burchett H, Umoquit M, Dobrow M. How do we know when research from one setting can be useful in another? A review of external validity, applicability and transferability frameworks. J Health Serv Res Policy 2011;16(4):238–44. doi: 10.1258/jhsrp.2011.010124

48. Smith GB, Redfern O, Maruotti A, et al. The association between nurse staffing levels and a failure to respond to patients with deranged physiology: A retrospective observational study in the UK. Resuscitation 2020 doi: https://doi.org/10.1016/j.resuscitation.2020.01.001

49. Griffiths P, Ball J, Bloor K, et al. Nurse staffing levels, missed vital signs and mortality in hospitals: retrospective longitudinal observational study. Health Services and Delivery Research Journal 2018;6(38) doi: 10.3310/hsdr06380

50. Redfern OC, Griffiths P, Maruotti A, et al. The association between nurse staffing levels and the timeliness of vital signs monitoring: a retrospective observational study in the UK. BMJ Open 2019;9(9):e032157. doi: 10.1136/bmjopen-2019-032157 [published Online First: 2019/09/29]

51. Ball JE, Bruyneel L, Aiken LH, et al. Post-operative mortality, missed care and nurse staffing in nine countries: A cross-sectional study. Int J Nurs Stud 2018;78:10–15. doi: 10.1016/j.ijnurstu.2017.08.004 [published Online First: 2017/08/29]

52. Twigg DE, Gelder L, Myers H. The impact of understaffed shifts on nurse-sensitive outcomes. J Adv Nurs 2015;71(7):1564–72.

53. Griffiths P, Jones, S., Maben, J., Murrels, T. State of the art metrics for nursing: a rapid appraisal: National Nursing Research Unit, 2008.

54. Blume KS, Dietermann K, Kirchner-Heklau U, et al. Staffing levels and nursing-sensitive patient outcomes: Umbrella review and qualitative study. Health Serv Res 2021;n/a(n/a) doi: 10.1111/1475-6773.13647 [published Online First: 2021/03/17]

