## Appendix 1 for "Nurse staffing levels and patient outcomes: a systematic review of longitudinal studies"

**MEDLINE**

| **ID#** | **Search Term** | **Results** |
| --- | --- | --- |
| 1 | staffing.mp. | 20,661 |
| 2 | exp Workforce/ | 44,128 |
| 3 | exp "Personnel Staffing and Scheduling"/ | 35,711 |
| 4 | (skill* adj1 mix*).tw. | 890 |
| 5 | (staff* adj3 (level* or ratio* or model* or roster*)).mp. | 5,117 |
| 6 | exp Workload/ | 20,444 |
| 7 | workload.mp. | 34,476 |
| 8 | 1 or 2 or 3 or 4 or 5 or 6 or 7 | 97,525 |
| 9 | nurs*.mp. | 430,548 |
| 10 | Nursing Staff, Hospital/ | 29,188 |
| 11 | Nursing Services/ | 966 |
| 12 | exp Nursing Assistants/ | 2,765 |
| 13 | ("health care assistant*" or "healthcare assistant*").mp. | 649 |
| 14 | ("health care support worker*" or "healthcare support worker*").mp. | 71 |
| 15 | "midwi*".mp. [mp=title, abstract, original title, name of substance word, subject heading word, floating sub-heading word, keyword heading word, organism supplementary concept word, protocol supplementary concept word, rare disease supplementary concept word, unique identifier, synonyms] | 23,703 |
| 16 | exp Midwifery/ | 13,491 |
| 17 | 9 or 10 or 11 or 12 or 13 or 14 or 15 or 16 | 439,126 |
| 18 | longitudinal.mp. | 248,836 |
| 19 | exp Longitudinal Studies/ | 130,948 |
| 20 | "cohort stud*".mp. | 389,847 |
| 21 | exp Cohort Studies/ | 1,851,439 |
| 22 | "panel stud*".mp. | 2159 |
| 23 | "prospective stud*".mp. | 557,198 |
| 24 | exp Prospective Studies/ | 511,107 |
| 25 | "randomi* control trial".mp. | 4698 |
| 26 | Randomized Controlled Trials as Topic/ | 138,567 |
| 27 | RCT.mp. | 21,107 |
| 28 | (cluster adj3 trial).tw. | 8658 |
| 29 | exp Survival Analysis/ | 302,442 |
| 30 | "survival analysis".mp. | 150,049 |
| 31 | 18 or 19 or 20 or 21 or 22 or 23 or 24 or 25 or 26 or 27 or 28 or 29 or 30 | 2,288,443 |
| 32 | 8 and 17 and 31 | 2336 |

**CINAHL**

| **ID#** | **Search Term** | **Results** |
| --- | --- | --- |
| 1 | (MH "Personnel Staffing and Scheduling+") | 33,593 |
| 2 | "staffing" | 31,182 |
| 3 | skill* N4 mix | 2,658 |
| 4 | staff* N3 (level* OR ratio* OR model* OR roster*) | 7,294 |
| 5 | "nursing hour* per patient day" | 74 |
| 6 | NHPPD | 20 |
| 7 | workload | 26,360 |
| 8 | (MH "Workload") | 16,634 |
| 9 | S1 OR S2 OR S3 OR S4 OR S5 OR S6 OR S7 OR S8 | 66,816 |
| 10 | (MH "Nurses+") | 238,428 |
| 11 | nurs* | 976,084 |
| 12 | (MH "Nursing Assistants") | 8,729 |
| 13 | (MH "RN First Assistants") | 859 |
| 14 | "nurs* aide*" | 853 |
| 15 | "healthcare assistant*" | 1,286 |
| 16 | "health care assistant*" | 554 |
| 17 | "healthcare support worker*" | 185 |
| 18 | "health care support worker*" | 56 |
| 19 | midwi* | 66,066 |
| 20 | (MH "Midwives+") | 16,148 |
| 21 | (MH "Midwifery Service+") | 2,027 |
| 22 | S10 OR S11 OR S12 OR S14 OR S15 OR S16 OR S17 OR S18 OR S19 OR S20 OR S21 | 1,014,023 |
| 23 | "longitudinal" | 95,410 |
| 24 | (MH "Prospective Studies+") | 478,531 |
| 25 | "panel stud*" | 2,365 |
| 26 | "cohort stud*" | 105,497 |
| 27 | "RCT" | 10,923 |
| 28 | cluster N2 trial | 7,352 |
| 29 | random* N3 trial | 244,626 |
| 30 | (MH "Randomized Controlled Trials+") | 118,475 |
| 31 | (MH "Survival Analysis+") | 93,468 |
| 32 | "survival analysis" | 40,060 |
| 33 | S23 OR S24 OR S25 OR S26 OR S27 OR S28 OR S29 OR S30 OR S31 OR S32 | 812,237 |
| 34 | S9 AND S22 AND S33 | 1,594 |

**COCHRANE LIBRARY**

| **ID#** | **Search Term** | **Results** |
| --- | --- | --- |
| 1 | MeSH descriptor: [Workforce] explode all trees | 420 |
| 2 | staffing | 725 |
| 3 | MeSH descriptor: [Personnel Staffing and Scheduling] explode all trees | 613 |
| 4 | MeSH descriptor: [Nursing] explode all trees | 3,311 |
| 5 | nurs* | 58,916 |
| 6 | MeSH descriptor: [Midwifery] explode all trees | 332 |
| 7 | midwi* | 5,410 |
| 8 | MeSH descriptor: [Longitudinal Studies] | 6,221 |
| 9 | longitudinal* | 22,822 |
| 10 | MeSH descriptor: [Prospective Studies] explode all trees | 93,296 |
| 11 | prospective | 237,426 |
| 12 | #1 OR #2 OR #3 | 1611 |
| 13 | #4 OR #5 OR #6 OR #7 | 61,036 |
| 14 | #8 OR #9 OR #10 OR #11 | 255,249 |
| 15 | #12 AND #13 AND #14 | 142 |
