## Appendix 2 for "Nurse staffing levels and patient outcomes: a systematic review of longitudinal studies"

| **Author(s), year** | **Country & setting** | **Design & Data collection period** | **Sample size** | **Data source** | **Outcomes & risk-adjustment** | **Domains affected by critical risk of bias** |
| --- | --- | --- | --- | --- | --- | --- |
| Ambrosi et al, 2017 | Italy, Acute Medical Units | Secondary analysis of prospective observational; 7 months: 2011-2012 | Hospitals = 12; AMUs =12; RNs=205; NAs =109; Patients = 1464 | Outcomes: unclear, but presumably routine administrative data. Nurses: staffing data collected by researchers daily. Risk-adjustment factors: a mixture of routine data and data collected specifically for project at bedside. | In-hospital mortality in a particular age group (>65 years); risk-adjusted for admission variables (age, recent A&E department visits, number of co-morbidities, admission type, admitted from home/nursing home, number of care problems/devices, behaviour disturbances, agitated/confused, BRASS Index score, at risk of pressure sores, Barthel Index score), during hospitalization variables (number of confusion/agitation events, pressure ulcers yes/no, physical restraints, care from family carers), discharge variables (functionally declined vs stable/improved, day of death), missed nursing care. | - **Confounding**   No information on diagnostic groups. Controlled for variables that should be mediators (i.e. missed care) and that are only applicable to patients who experienced the outcome (i.e. day of death)   - **Classification of interventions**   The classification of the intervention (i.e. receiving care from RN -12 to -24 min/day at weekend vs weekday compared to those receiving from -4 to -11 min/day) is not justified and/or rationalised |
| Jansson et al, 2019 | Finland, Medical / Surgical Intensive Care | Cohort; October 2014-June 2015 (8 months) | Hospitals= 1; Wards= 1; Patients= 85 | **Outcomes:**  Clinical databases**;**  **Staffing levels:** administrative data | **Primary outcome:** Ventilator associated pneumonia  **Secondary outcome:** 28-day mortality  No risk adjustment | - **Confounding:**   No risk-adjustment   - **Classification of interventions:**   Intervention groups were clearly defined for one outcome (VAP) but not for the secondary outcome (mortality).  The exposure window was defined at the start of exposure only for one outcome (VAP), but not for the secondary outcome (mortality) |
| Mark & Belyea, 2009 | US; acute care hospitals | Observational; January 2004 to June 2004 (6 months) | Hospitals= 145; Wards= 284 Nurses = 4911 | **Outcome data:**  Study coordinators  **Hospital data:** AHA Annual Survey  of Hospitals (2004);  **Ward data:**  On-site study coordinators; **Staffing data:** Onsite study coordinators | Medication errors, including wrong patient, drug, dose, time or route. Adjusted for: hospital environment (size, teaching status), unit environment (size, occupancy, work uncertainty, medication support systems) and patient workload (case mix, patient acuity) | - **Bias due to deviations from intended interventions:**   It cannot be established whether there were any deviations from the intended intervention beyond what would be expected in usual practice because the relationship between staffing levels and the outcome has one month lag, meaning that the staffing levels of month 1 could not be those influencing the patient outcome in month 2. |
| Palese et al, 2015 | Italy; Acute medical units | Observational; Longitudinal phase: January 2012 to March 2012 (3 months)cross-sectional phase: March 2012 (1 month) | Hospitals= 12; Wards= 12 Registered Nurses= 205; Nursing Aides=109 | **Staffing data:** direct observation by two trained researchers;  **Outcome:** (cross-sectional phase): MISSCARE survey tool  **Staff demographic data:** open-ended questionnaire | Missed nursing care as defined in MISSCARE (24 nursing interventions)  Risk-adjusted for working position (full-time versus part-time), communicative tensions between RNs and NAs, experience on the unit, staff age | - **Bias in measurement of outcomes:**   The outcome measure may have been influenced by knowledge of the intervention received, since nurses were asked to report their missed care as part of a study about staffing levels. It is possible that nurses who thought staffing levels were too low would indicate higher rates of missed care.  Outcome assessors were aware of the intervention received by study participants, since nurses who filled in a MISSCARE survey knew which staffing levels they had been exposed to.   - There were errors in measurement of the outcome related to intervention received; there was an incorrect conceptualisation of missed care outcome: the Likert-type scale was dichotomized into no missed care (Likert scale from 1 to 2, never or rarely missed care) and a positive response of missed care (Likert scale from 3 to 5, occasionally, frequently, or always missed care). However, a “rarely” missed care categorisation is not “no missed care” |
| Palese et al, 2016 | Italy; Acute Medical Units | Prospective observational; 2012-2013 | Hospitals =12; AMUs =12; RNs= 252; NAs = 165; patients = 1464 | Outcome: Barthel Index score at discharge measured by Research Nurses . Nurses: daily Chief nurse interviews and MISSCARE survey of nurses. Risk-adjustment: admin records, observations and patient/caregiver interviews. | Functional decline in a particular age group (>65 years), measured as a decrease in the Barthel Index score of at least five points from admission to discharge; risk-adjusted for patient variables (age, gender, length of stay, confused/disoriented, agitated/wondered, recent admission to A&E, number of medications, number of co-morbidities, impairments in functional status before admission, bladder catheter insertion at admission, Barthel Index score at admission, admission for nursing home, % of days physically constrained, % of days with caregivers at bedside, falls), nursing variables (missed care) and hospital variables (size, highly specialized). | - **Confounding:**   No information on diagnostic groups. Controlled for variables that should be mediators (i.e. missed care) and no ward-level effects adjustment/no inclusion of wards as random effects. Due to moderator being treated as a control variable, we do not know whether the moderator may increase the strength of the relationship, or change the direction of the relationship |
| Twigg et al, 2016 | Australia, acute care hospitals | Pre-test/post-test; data collected in pre-test (2006-2007) and post-test (2009-2010) | Hospitals = 11; Wards =64;  Patients = 125,762 pre-test, 130,540 in post-test | **Patient outcomes:** Routinely collected dataset obtained through WA Data Linkage Unit  **Nurse staffing:**  Not specified | 30-day mortality; failure to rescue; urinary tract infections; pressure injury; pneumonia; sepsis; falls with injury.  Adjusted for: patient group (medical or surgical admission), age, gender, season (of admission), Indigenous status, source of referral to the hospital (home, nursing home/hostel, other hospital, other), Diagnosis Related Group (DRG) cost weight (a measure of the resource use of each patient), age squared, peer group of the hospital (type of hospital), length of stay, elective or emergency admission, Charlson comorbidity index, and DRG cost weight by age, and gender by age interactions. | - **Selection of the reported result:**   There were multiple analyses of the association between staffing levels and outcomes. In Table 3 authors report two ‘before and after’ studies (effect of adding AIN and the effect of not adding AIN) - analytically this is not a before and after study and so it is (effectively) uninterpretable as it stands. In addition, the effect measure is also uninterpretable because it is impossible to establish when is observed (?after) and what gives expected. |
