## Appendix 3 for "Nurse staffing levels and patient outcomes: a systematic review of longitudinal studies"

Appendix 3 Risk of bias assessment by domain and risk of bias direction

| **Author(s), year** | **Confounding** | **Participants** | **interventions** | **Deviations** | **Missing data** | **Outcomes** | **Results** | **Overall bias** | **Risk of bias direction** |
| --- | --- | --- | --- | --- | --- | --- | --- | --- | --- |
| Al-Abdwani et al, 2018 | Serious | Low | Serious | Low | Low | Low | Low | Serious | Favours lower staffing levels |
| Alonso-Echanove, 2003 | Serious | Low | Moderate | Serious | Low | Serious | Serious | Serious | Favours higher (float nurse) staffing levels |
| Ambrosi et al, 2017 | Critical | Low | Serious | Low | Low | Low | Critical | Critical | Unpredictable |
| Beltempo et al, 2017 | Serious | Low | Moderate | Low | Low | Low | Moderate | Serious | Favours lower staffing levels |
| Beltempo et al, 2018 | Moderate | Moderate | Moderate | Low | Low | Low | Moderate | Moderate | Unpredictable |
| Callaghan et al, 2003 | Serious | Moderate | Moderate | Low | Low | Low | Moderate | Serious | Favours lower staffing levels |
| Cimiotti et al, 2004; Cimiotti et al 2006 | Serious | Low | Low | Low | Low | Low | Serious | Serious | Favours lower staffing levels |
| Dall’Ora et al, 2020 | Serious | Low | Moderate | Low | Low | Low | Moderate | Serious | Unpredictable |
| Fogg et al, 2021 | Moderate | Low | Low | Low | Low | Low | Moderate | Moderate | Unpredictable |
| Griffiths et al, 2018 | Low | Low | Low | Low | Low | Low | Moderate | Moderate | Unpredictable |
| Griffiths et al, 2019 | Low | Low | Low | Low | Low | Low | Low | Low |  |
| Hamilton et al, 2007 | Moderate | Low | Low | Low | Moderate | Moderate | Moderate | Moderate | Favours higher staffing levels |
| Hugonnet et al, 2007a - staffing levels | Serious | Low | Moderate | Low | Low | Moderate | Moderate | Serious | Favours lower staffing levels |
| Hugonnet et al, 2007b - workload | Serious | Low | Moderate | Low | Low | Low | Serious | Serious | Favours lower staffing levels |
| Jansson et al, 2019 | Critical | Low | Critical | Low | Low | Low | Serious | Critical | Favours lower staffing levels |
| Mark & Belyea, 2009 | Serious | Low | Serious | Critical | Moderate | Low | Serious | Critical | Unpredictable |
| McHugh et al, 2021 | Moderate | Serious | Moderate | Low | Low | Low | Serious | Serious | Unpredictable |
| Musy et al, 2021 | Low | Low | Low | Low | Low | Low | Low | Low |  |
| Needleman et al, 2011 | Moderate | Low | Low | Low | Low | Low | Moderate | Moderate | Unpredictable |
| Needleman et al, 2020 | Low | Low | Low | Low | Low | Low | Moderate | Moderate | Unpredictable |
| Palese et al, 2015 | Serious | Moderate | Moderate | Low | Low | Critical | Serious | Critical | Favours higher staffing levels |
| Palese et al, 2016 | Critical | Moderate | Low | Low | Low | Moderate | Moderate | Critical | Unpredictable |
| Patrician et al, 2017 | Serious | Moderate | Moderate | Low | Serious | Low | Serious | Serious | Favours lower staffing levels |
| Rochefort et al, 2020 | Low | Low | Low | Low | Low | Low | Low | Low |  |
| Shang et al, 2019 | Moderate | Low | Low | Low | Low | Low | Moderate | Moderate | Favours higher staffing levels |
| Tschannen et al, 2005 | Serious | Moderate | Moderate | Low | Low | Low | Moderate | Serious | Favours lower staffing levels |
| Twigg et al, 2016 | Moderate | Serious | Low | Low | Low | Low | Critical | Critical | Unpredictable |
